# Baseline substrate and response after cardiac resynchronization therapy in non-left bundle branch block heart failure

**DOI:** 10.64898/2026.05.14.26353260

**Authors:** Yueying Liang, Ye Zhu, Ruotong Wang, Renjie Gu, Chuanyi Sang, Zhenyu Bao, Lei Sun, Tian Xia, Xiang Gu

## Abstract

**Background:** Response to cardiac resynchronization therapy (CRT) is heterogeneous in patients with non-left bundle branch block (non-LBBB) heart failure. Whether pre-implant substrate or procedural characteristics provide the more stable framework for predicting 1-year echocardiographic response remains uncertain.

**Methods:** We retrospectively analyzed 120 non-LBBB patients undergoing CRT. The primary logistic model included left ventricular end-diastolic diameter (LVEDD), left ventricular ejection fraction (LVEF), left atrial diameter, log-transformed NT-proBNP, baseline QRS duration, fragmented QRS burden across V1–V6 leads, and pulmonary artery pressure. Missing predictor data were handled using multiple imputation with 20 datasets. Model performance was assessed using bootstrap internal validation and recalibration. A prespecified procedural extension added pacing strategy, posterolateral biventricular left ventricular lead location, left ventricular pacing threshold, and right ventricular lead position. Exploratory phenotyping and sensitivity analyses were performed.

**Results:** Echocardiographic response occurred in 51 patients (42.5%). LVEDD (OR, 0.899 [95% CI, 0.826–0.978]; P=0.013) and LVEF (OR, 1.068 [95% CI, 1.000– 1.140]; P=0.050) were the most informative predictors. The primary model showed apparent AUC 0.811 and Brier score 0.173, with optimism-corrected AUC 0.766 and calibration slope 0.765. Procedural extension showed no retained incremental value after validation. Exploratory phenotyping identified three response patterns with moderate stability.

**Conclusions:** In non-LBBB CRT, baseline structural, biomarker, and electrocardiographic substrate provided the most stable framework for predicting 1-year echocardiographic response. Procedural variables added limited retained value, suggesting that pacing strategy should be interpreted alongside baseline substrate.

## Introduction

Cardiac resynchronization therapy (CRT) is an established treatment for selected patients with heart failure and ventricular conduction delay, with the most consistent benefit observed in patients with typical left bundle branch block (LBBB)^[1-6]^. Contemporary guidelines emphasize QRS morphology, QRS duration, and left ventricular dysfunction when selecting candidates for CRT. However, patients with non-left bundle branch block (non-LBBB) conduction remain a heterogeneous and less predictable population, in whom echocardiographic response is less reliably anticipated by conventional eligibility criteria alone.

This uncertainty is partly explained by the fact that non-LBBB is an umbrella electrocardiographic category rather than a uniform substrate for resynchronization^[7,8]^. Patient-level evidence from randomized CRT trials suggests that treatment effects differ across non-LBBB morphologies: CRT was associated with improved outcomes in patients with intraventricular conduction delay and wide QRS, but not in those with right bundle branch block, challenging the practice of grouping these patterns under a single non-LBBB label. This distinction is clinically important because a wide QRS complex outside typical LBBB does not necessarily indicate the same pattern of delayed left ventricular activation, mechanical dyssynchrony, or reversible myocardial substrate that underlies the most predictable response to conventional biventricular CRT.

The therapeutic landscape of resynchronization has also changed with the emergence of conduction system pacing, particularly left bundle branch area pacing (LBBAP), as a physiologically attractive alternative or complement to conventional biventricular pacing^[9-14]^.Observational and comparative studies have suggested potential advantages of LBBAP in selected CRT candidates, whereas other contemporary work has highlighted the need for individualized procedural strategies, particularly in patients without typical LBBB.These developments raise an important clinical question: in non-LBBB patients, should response be interpreted primarily through procedural strategy, or through the baseline structural and electrical substrate in which that strategy is applied?

A substrate-based approach may help address this question. CRT response is increasingly understood as a multidimensional phenomenon shaped by ventricular remodeling, systolic function, atrial and hemodynamic burden, biomarker profile, electrical delay, and myocardial activation patterns rather than by a single electrocardiographic label^[17,18,21,22]^.Predictive modeling and phenotyping studies have suggested that multidimensional features can improve response characterization and identify clinically interpretable phenogroups among CRT candidates.However, few studies have focused specifically on non-LBBB recipients while also evaluating whether procedural variables add stable predictive information beyond pre-implant substrate.

In this study, we developed and internally validated a pre-implant substrate-based model for 1-year echocardiographic CRT response in patients with non-LBBB conduction. We further evaluated whether a prespecified procedural extension incorporating pacing strategy, LV lead location, LV pacing threshold, and RV lead position added incremental predictive value beyond baseline substrate. Finally, we performed exploratory phenotype analysis to characterize response heterogeneity within the non-LBBB cohort and to generate hypotheses for future phenotype-guided CRT studies.

## Methods

### Study Design and Population

This single-center retrospective cohort study screened consecutive patients undergoing CRT implantation, including conventional biventricular pacing and left bundle branch area pacing strategies, at our institution between January 2019 and December 2024. Eligible patients were adults with symptomatic heart failure, non-left bundle branch block (non-LBBB) conduction on the pre-implant electrocardiogram, clinically indicated CRT implantation, source records containing baseline clinical, electrocardiographic, laboratory, echocardiographic, and procedural information, and follow-up echocardiography sufficient to determine echocardiographic CRT response. Patients with LBBB morphology, indeterminate conduction morphology, unsuccessful or aborted CRT delivery, missing follow-up echocardiography, duplicate records, or unverifiable data were excluded. The detailed cohort derivation is shown in Figure 1. The study was approved by the Ethics Committee of Northern Jiangsu People’s Hospital, Yangzhou, China (approval number: 2021ky007), and the requirement for written informed consent was waived because of the retrospective design and use of de-identified clinical data.

**Figure 1.**
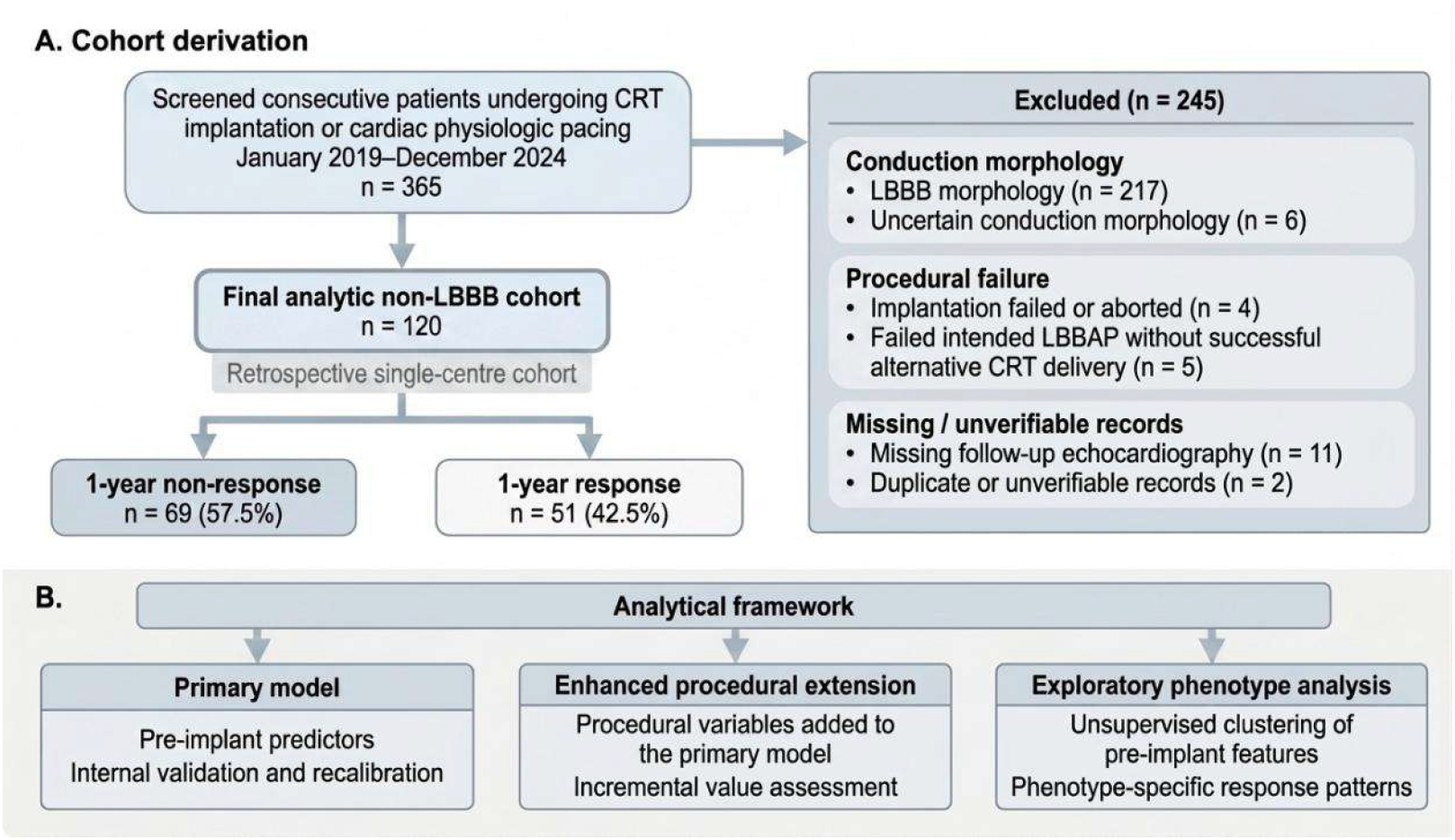
Study cohort derivation and analytical framework. Panel A shows derivation of the study cohort. Panel B summarizes the three analytical components of the study.

### Outcome Definition

The primary endpoint was 1-year echocardiographic CRT response, defined as an absolute increase in left ventricular ejection fraction (LVEF) of ≥10 percentage points from baseline. Patients meeting this criterion were classified as responders, whereas the remaining patients were classified as non-responders. Change in left ventricular end-diastolic diameter (LVEDD) from baseline to follow-up was additionally evaluated as a secondary echocardiographic remodeling measure, but was not used as a predictor in the primary model.

### Candidate Predictors

Pre-implant echocardiographic variables included LVEDD, LVEF, left atrial diameter (LAD), and pulmonary artery pressure (PAP), all measured on baseline transthoracic echocardiography before CRT implantation. The biomarker variable was pre-implant NT-proBNP, which was log-transformed before analysis because of its skewed distribution. Electrical variables included baseline QRS duration and fragmented QRS burden. Baseline QRS duration was measured immediately before implantation using the intra-procedural multi-lead electrophysiology recording system under intrinsic conduction, before delivery of CRT pacing. Fragmented QRS burden was assessed on the pre-implant 12-lead surface electrocardiogram and summarized as Vfrag count, defined as the number of precordial leads from V1 to V6 showing notched or fragmented QRS morphology.

### Procedural Variables

Procedural variables were recorded from implantation reports and peri-implant device testing. Device type, pacing strategy, LV pacing threshold, RV lead position, and BiV LV lead location were collected for descriptive and sensitivity analyses. Pacing strategy was classified as left bundle branch area pacing (LBBAP) or biventricular pacing (BiV). For BiV pacing, LV lead location was classified from procedural fluoroscopic documentation as lateral, posterolateral, or anterior/anterolateral; lateral location was used as the reference category for BiV lead-location analyses. RV lead position was classified as septal or apical. In the prespecified enhanced model, procedural extension variables were limited to pacing strategy, posterolateral BiV LV lead location relative to lateral BiV LV lead location, LV pacing threshold, and RV lead position. Additional procedural adjudication details are provided in the Supplementary Methods.

### Missing Data

Missing data in candidate predictors were addressed using multiple imputation with 20 imputed datasets. The imputation model included the outcome as a predictor, together with the prespecified primary-model predictors, procedural extension variables, and variables required for sensitivity analyses; however, the outcome itself was not imputed. Post-treatment variables were excluded to avoid information leakage. Imputation was performed using IterativeImputer with posterior sampling. Binary variables were constrained to valid 0/1 values after imputation, and Vfrag count was restricted to the allowable range of 0–6. Temporal variables were retained for sensitivity analyses but were not imputed. Complete-case analysis was performed as a prespecified sensitivity analysis^[23]^.

### Model Development and Validation

The primary model was developed using logistic regression with a prespecified set of pre-implant predictors: LVEDD, LVEF, LAD, log-transformed NT-proBNP, baseline QRS duration, Vfrag count, and PAP. Model performance was assessed by discrimination, overall accuracy, and calibration, using the area under the receiver operating characteristic curve, Brier score, calibration intercept, and calibration slope. Internal validation was performed using 500 bootstrap resamples to estimate optimism-corrected performance. Recalibration strategies, including intercept recalibration and global shrinkage, were evaluated, and the intercept-recalibrated primary model was selected as the preferred version for main-text reporting. The enhanced model was prespecified as a procedural extension of the primary model by adding pacing strategy, posterolateral BiV LV lead location, LV pacing threshold, and RV lead position. Its incremental value was evaluated by comparing apparent and optimism-corrected performance against the primary model^[24,25]^.

### Sensitivity Analyses

Sensitivity analyses compared complete-case and multiply imputed analyses, replaced Vfrag count with individual V1–V6 fragmentation indicators, removed the BiV lead-position indicator from the enhanced model, compared recalibration strategies, and explored temporal/era effects descriptively. These analyses were prespecified and were not used to redefine the primary model.

### Exploratory Phenotype Analysis

Exploratory phenotype analysis was conducted using six pre-implant variables: LVEDD, LVEF, LAD, log-transformed NT-proBNP, baseline QRS duration, and Vfrag count. The outcome and procedural variables were not used for clustering. Continuous variables were standardized before clustering. Candidate cluster solutions with k=2, k=3, and k=4 were evaluated using clustering performance metrics and clinical interpretability. Cluster labels were aligned across imputed datasets, and cluster stability was assessed using the adjusted Rand index. Echocardiographic response rates were compared only after phenotype group assignment. These analyses were considered exploratory and hypothesis-generating.

### Statistical Analysis

Continuous variables were reported as mean±SD or median [interquartile range], and categorical variables as counts and percentages. Standardized differences were used to describe between-group imbalance. Estimates from the imputed datasets were pooled using Rubin’s rules. Model performance was evaluated using the area under the receiver operating characteristic curve (AUC), Brier score, calibration intercept, and calibration slope, with calibration plots used for graphical assessment. Data quality checks included range assessment, logical consistency checks, and verification of categorical levels before and after imputation. P values were two-sided and interpreted descriptively. Sensitivity analyses and exploratory phenotype analyses were interpreted descriptively and did not alter the prespecified primary model. Analyses were performed using Python with pandas, numpy, scikit-learn, statsmodels, scipy, and matplotlib. Model reporting and interpretation were guided by contemporary recommendations for clinical prediction model studies^[26,27]^.

## Results

### Study population and baseline characteristics

Between January 2019 and December 2024, 365 consecutive patients undergoing CRT implantation, including conventional biventricular pacing and left bundle branch area pacing strategies, were screened. After excluding patients with LBBB morphology (n=217), indeterminate conduction morphology (n=6), unsuccessful or aborted CRT delivery (n=9, including 5 patients with failed intended left bundle branch area pacing without successful alternative CRT delivery), missing follow-up echocardiography (n=11), or duplicate/unverifiable records (n=2), 120 patients with non-LBBB conduction formed the final analytic cohort (Figure 1). Echocardiographic CRT response at 1 year was observed in 51 patients (42.5%), whereas 69 patients (57.5%) were classified as nonresponders.

Baseline characteristics of the overall cohort and according to response status are shown in Table 1. The cohort was elderly and showed substantial baseline structural remodeling, with a mean LVEDD of 57.9 mm and mean LVEF of 42.2%. Compared with nonresponders, responders had smaller LVEDD, higher LVEF, smaller LAD, and lower mitral regurgitation grade, consistent with a more favorable baseline structural profile. Responders also had lower log NT-proBNP and narrower baseline QRS duration, whereas the distribution of non-LBBB subtypes was broadly similar between groups. Procedural characteristics, including pacing strategy, BiV LV lead location, RV lead position, and LV pacing threshold, are also summarized in Table 1.

**Table 1.**
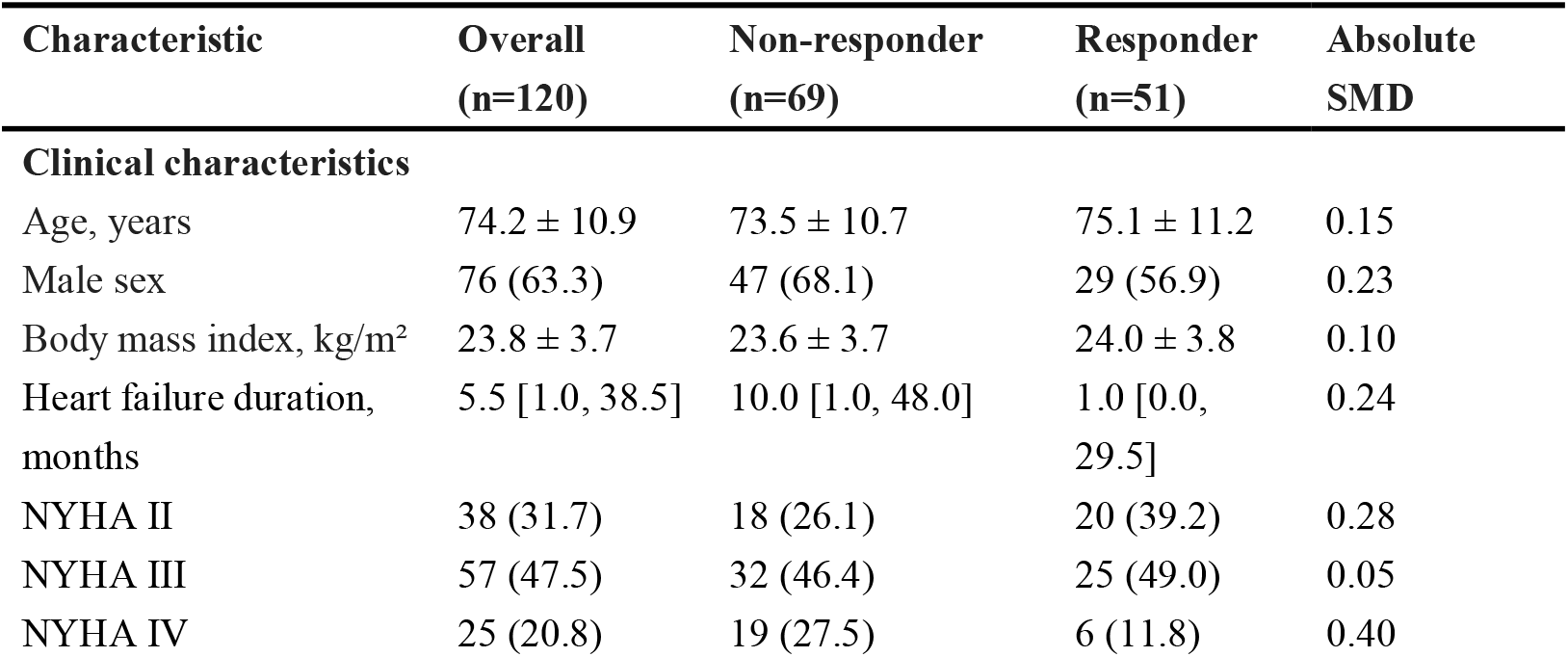
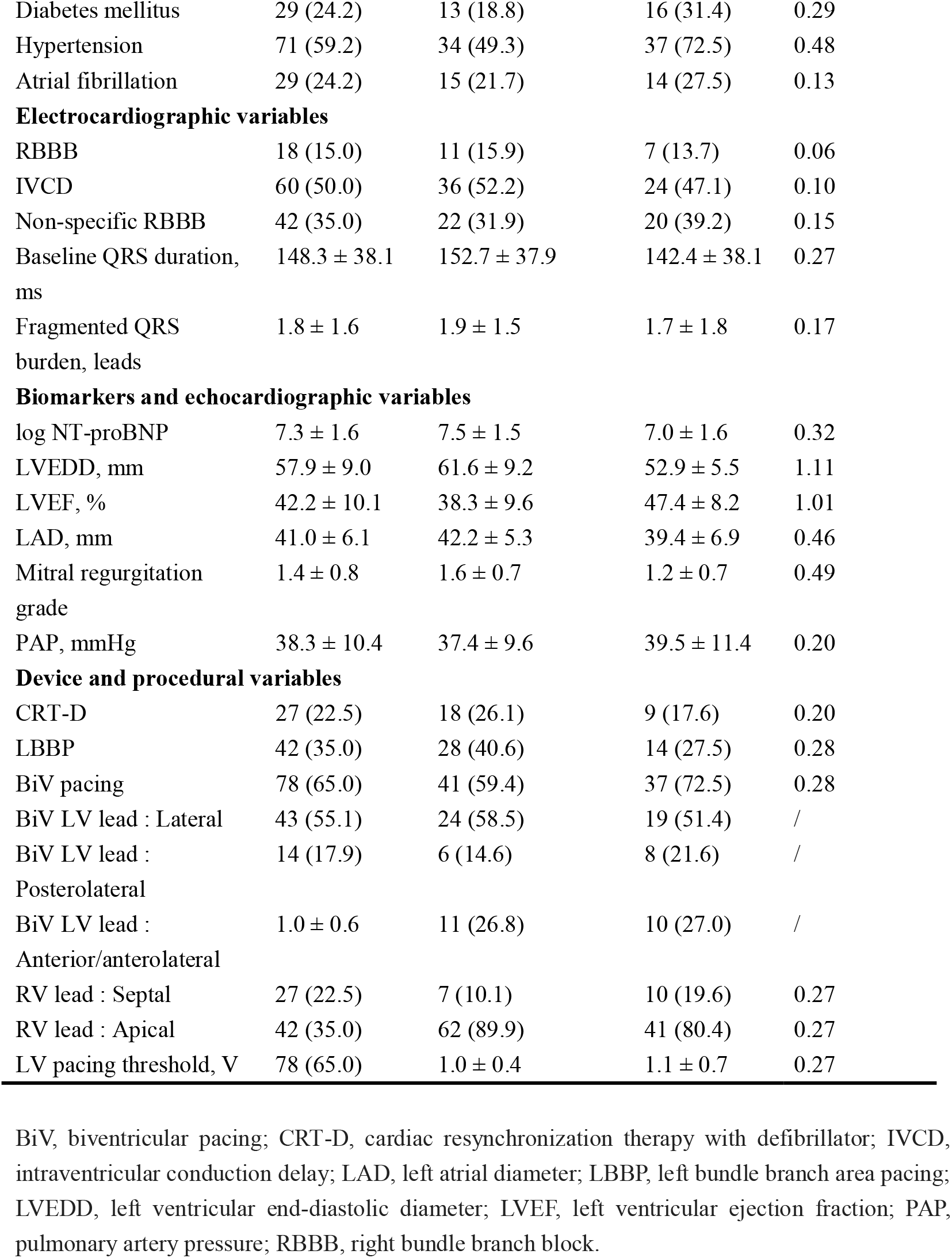
Baseline characteristics of the overall cohort and stratified by echocardiographic response status.

### Primary model performance and internal validation

The primary model incorporated seven pre-implant predictors: LVEDD, LVEF, LAD, log NT-proBNP, baseline QRS duration, Vfrag count, and PAP. In the pooled regression analysis, LVEDD and LVEF showed the strongest associations with echocardiographic CRT response. Each 1-mm increase in LVEDD was associated with lower odds of response (OR, 0.899 [95% CI, 0.826–0.978]; P=0.013), whereas each 1-percentage point increase in LVEF was associated with higher odds of response (OR, 1.068 [95% CI, 1.000–1.140]; P=0.050). The remaining predictors showed less precise associations (Table 2).

**Table 2.** Primary model coefficients, internal validation, and recalibration.

| **Panel A. Pooled coefficients of the primary model** |  |  |  |  |
| --- | --- | --- | --- | --- |
| Predictor | Coefficient | SE | OR (95% CI) | P value |
| LVEDD, per mm | -0.107 | 0.043 | 0.899 (0.826–0.978) | 0.013 |
| LVEF, per % | 0.065 | 0.033 | 1.068 (1.000–1.140) | 0.050 |
| LAD, per mm | -0.052 | 0.046 | 0.949 (0.867–1.040) | 0.264 |
| log NT-proBNP | 0.025 | 0.175 | 1.026 (0.727–1.446) | 0.885 |
| Baseline QRS duration, per ms | 0.006 | 0.007 | 1.006 (0.992–1.020) | 0.418 |
| Fragmented QRS burden, per lead | -0.244 | 0.164 | 0.784 (0.568–1.082) | 0.138 |
| PAP, per mmHg | 0.046 | 0.029 | 1.047 (0.989–1.107) | 0.112 |

**Panel B. Model performance and recalibration**
| Metric | Apparent | Optimism-corrected | Intercept-recalibrated |
| --- | --- | --- | --- |
| AUC | 0.811 | 0.766 | 0.820 |
| Brier score | 0.173 | 0.201 | 0.169 |
| Calibration intercept | 0.000 | -0.030 | 0.012 |
| Calibration slope | 1.000 | 0.765 | 1.104 |

Values in Panel A are pooled across 20 imputed datasets using Rubin’s rules. Optimism-corrected metrics were estimated by bootstrap internal validation. LVEDD, left ventricular end-diastolic diameter; LVEF, left ventricular ejection fraction; LAD, left atrial diameter; PAP, pulmonary artery pressure; OR, odds ratio; SE, standard error; AUC, area under the receiver operating characteristic curve.

In the multiply imputed datasets, the primary model showed an apparent AUC of 0.811 and a Brier score of 0.173. Apparent calibration metrics were close to ideal, with a calibration intercept of approximately 0 and calibration slope of approximately 1.0. Bootstrap internal validation attenuated model performance, yielding an optimism-corrected AUC of 0.766 and Brier score of 0.201, with a calibration intercept of −0.030 and calibration slope of 0.765 (Table 2). After recalibration, the intercept-recalibrated primary model showed an AUC of 0.820, Brier score of 0.169, calibration intercept of 0.012, and calibration slope of 1.104, and was retained as the preferred primary-model version (Figure 2).

**Figure 2.**
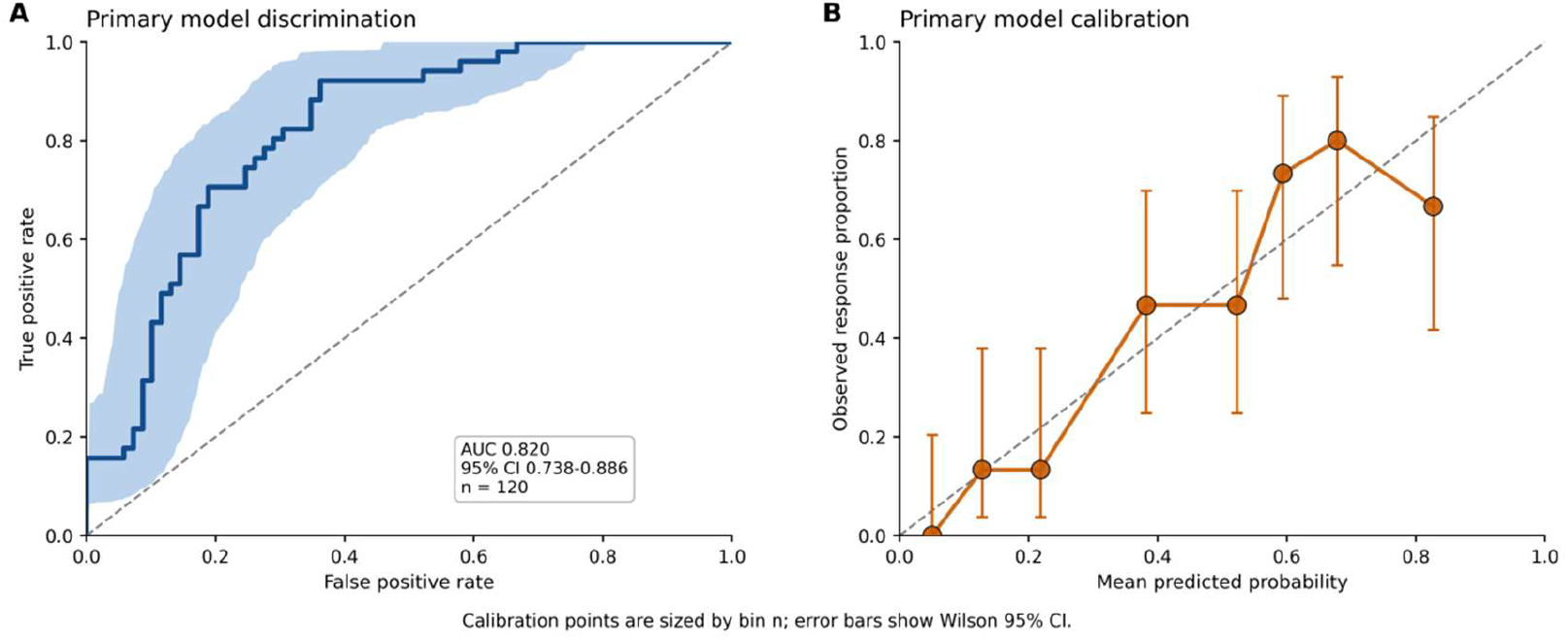
Discrimination and calibration of the primary phenotype-based model. (A) Receiver operating characteristic curve of the primary pre-implant phenotype-based model for predicting 1-year echocardiographic CRT response in patients with non-left bundle branch block (non-LBBB) conduction. (B) Calibration plot of the intercept-recalibrated primary model. Points represent grouped observed response rates plotted against mean predicted probabilities; the dashed line indicates ideal calibration.

### Procedural extension model

The procedural extension model incorporated all primary-model predictors and added pacing strategy, posterolateral BiV LV lead location relative to lateral BiV LV lead location, LV pacing threshold, and RV lead position. In the pooled enhanced model, LVEDD remained the most robust predictor of echocardiographic CRT response, with each 1-mm increase associated with lower odds of response (OR, 0.895 [95% CI, 0.819–0.979]; P=0.015), whereas LVEF showed a directionally consistent but less statistically robust association (OR, 1.064 [95% CI, 0.994–1.138]; P=0.072). The added procedural variables did not show statistically robust independent associations. Although the enhanced model showed a small apparent improvement over the primary model (AUC, 0.818 versus 0.811; ΔAUC, 0.007; Brier score, 0.170 versus 0.173), this gain was not retained after bootstrap internal validation. The optimism-corrected AUC was lower for the enhanced model than for the primary model (0.748 versus 0.766), and the corrected Brier score was higher (0.213 versus 0.201). Accordingly, the prespecified comparison supported retaining the primary model rather than replacing it with the procedural extension model (Table 3; Figure 3).

**Table 3.** Enhanced procedural extension model and comparison with the primary model.

**Panel A. Pooled coefficients of the enhanced procedural extension model**
| Predictor | Coefficient | SE | OR (95% CI) | P value |
| --- | --- | --- | --- | --- |
| LVEDD, per mm | -0.111 | 0.046 | 0.895 (0.819–0.979) | 0.015 |
| LVEF, per % | 0.062 | 0.034 | 1.064 (0.994–1.138) | 0.072 |
| LAD, per mm | -0.055 | 0.047 | 0.946 (0.863–1.038) | 0.241 |
| log NT-proBNP | 0.015 | 0.182 | 1.015 (0.710–1.450) | 0.935 |
| Baseline QRS duration, per ms | 0.007 | 0.007 | 1.007 (0.992–1.021) | 0.371 |
| Vfrag count, per fragmented lead | -0.256 | 0.164 | 0.774 (0.561–1.069) | 0.120 |
| PAP, per mmHg | 0.047 | 0.030 | 1.048 (0.988–1.112) | 0.121 |
| BiV pacing vs LBBP | 0.509 | 0.548 | 1.664 (0.568–4.869) | 0.353 |
| Posterolateral BiV LV lead location indicator | 0.475 | 0.761 | 1.608 (0.362–7.149) | 0.533 |
| LV pacing threshold | -0.101 | 0.407 | 0.904 (0.407–2.006) | 0.803 |
| RV lead position, apical vs septal | -0.221 | 0.656 | 0.802 (0.222–2.900) | 0.736 |

| **Panel B. Primary versus enhanced model performance** |  |  |  |  |
| --- | --- | --- | --- | --- |
| Performance level | Metric | Primary model | Enhanced model | Difference (enhanced - primary) |
| Apparent performance | AUC | 0.811 | 0.818 | 0.007 |
| Apparent performance | Brier score | 0.173 | 0.170 | -0.003 |
| Apparent performance | Calibration intercept | 0.000 | 0.000 | 0.000 |
| Apparent performance | Calibration slope | 1.000 | 1.000 | 0.000 |
| Optimism-corrected performance | AUC | 0.766 | 0.748 | -0.017 |
| Optimism-corrected performance | Brier score | 0.201 | 0.213 | 0.012 |
| Optimism-corrected performance | Calibration intercept | -0.030 | -0.049 | -0.020 |
| Optimism-corrected performance | Calibration slope | 0.765 | 0.649 | -0.117 |

Values in Panel A are pooled across 20 imputed datasets using Rubin's rules. The intercept is omitted from the table. The enhanced model was constructed as the primary model plus procedural extension variables. BiV, biventricular pacing; LBBP, left bundle branch area pacing; LVEDD, left ventricular end-diastolic diameter; LVEF, left ventricular ejection fraction; LAD, left atrial diameter; PAP, pulmonary artery pressure; OR, odds ratio; SE, standard error; AUC, area under the receiver operating characteristic curve. Posterolateral BiV LV lead location indicator denotes the position variable evaluated in the simplified enhanced model and should be interpreted conditional on pacing strategy. RV lead position was coded as apical versus septal according to the procedural record.

**Figure 3.**
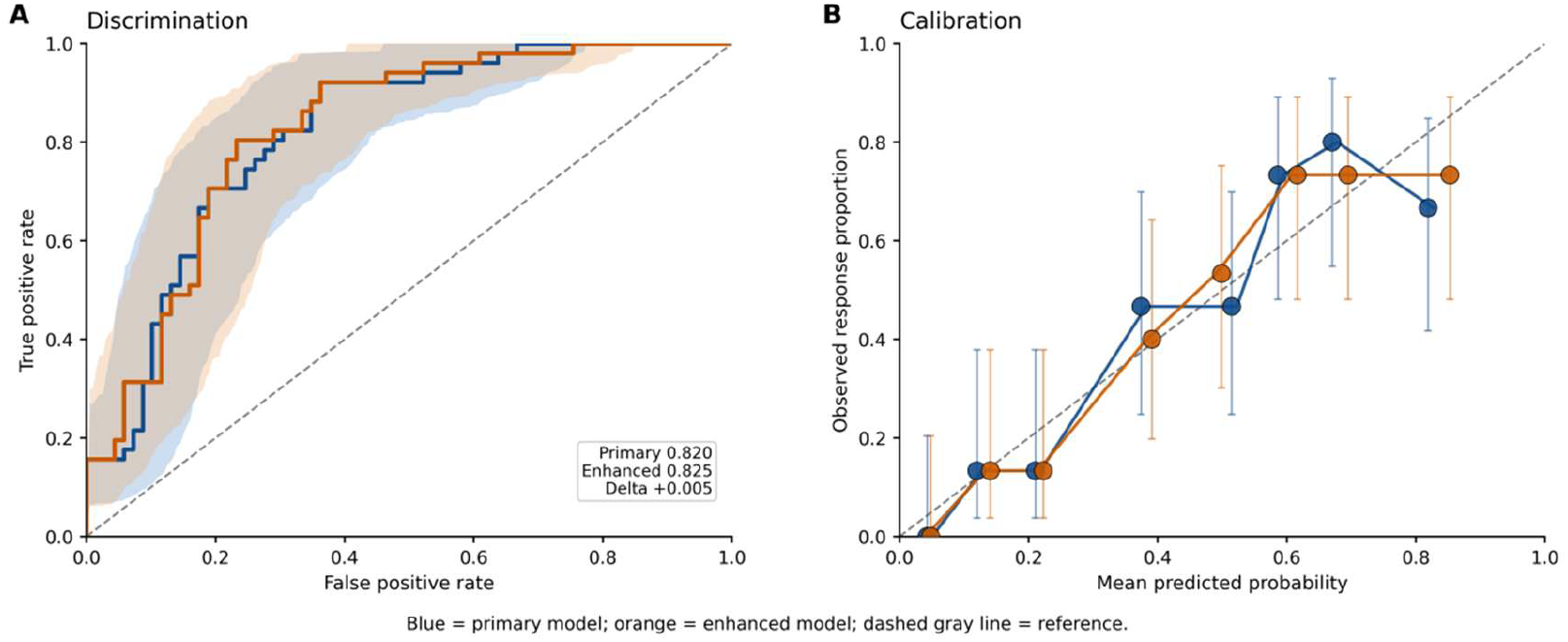
Comparison of the primary model and procedural extension model. (A) Receiver operating characteristic curves comparing the primary pre-implant phenotype-based model and the prespecified procedural extension model. (B) Calibration comparison between the primary model and the procedural extension model, with the dashed line indicating ideal calibration.

### Exploratory phenotype analysis

Exploratory unsupervised phenotyping identified a three-group solution based on prespecified pre-implant phenotype variables. Phenotype 1 comprised a mean of 36.8 patients across imputed datasets and showed a response rate of 53.2%. Phenotype 2 comprised a mean of 35.0 patients and had the highest response rate at 59.9%. Phenotype 3 was the largest group, comprising a mean of 48.2 patients, but had the lowest response rate at 22.0% (Table 4).

**Table 4.**
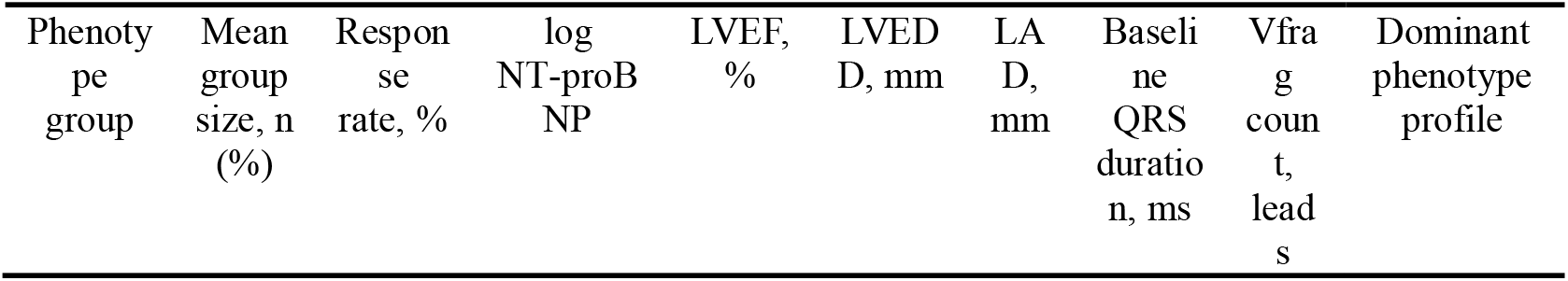
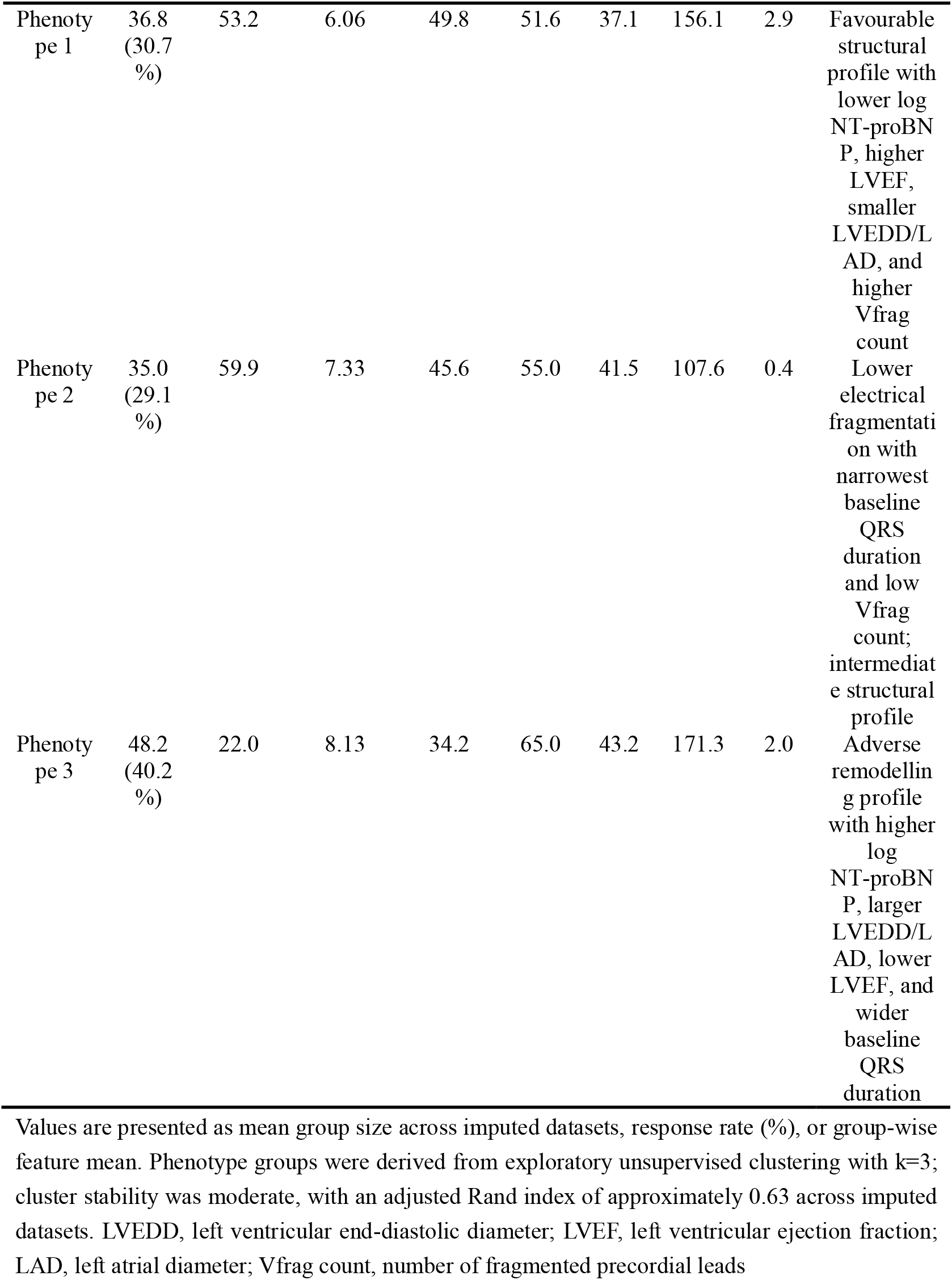
Exploratory phenotype groups and echocardiographic response rates.

The three groups differed in their structural, biomarker, and electrical profiles. Phenotype 1 showed a relatively favorable structural profile, with lower log NT-proBNP, higher LVEF, smaller LVEDD and LAD, and higher Vfrag count. Phenotype 2 was characterized by lower electrical fragmentation, the narrowest baseline QRS duration, and an intermediate structural profile. Phenotype 3 showed an adverse remodeling profile, with higher log NT-proBNP, larger LVEDD and LAD, lower LVEF, and wider baseline QRS duration. Cluster stability across imputed datasets was moderate, with an adjusted Rand index of approximately 0.63; these groups were therefore interpreted as exploratory phenotype patterns rather than definitive biological subtypes (Figure 4).

**Figure 4.**
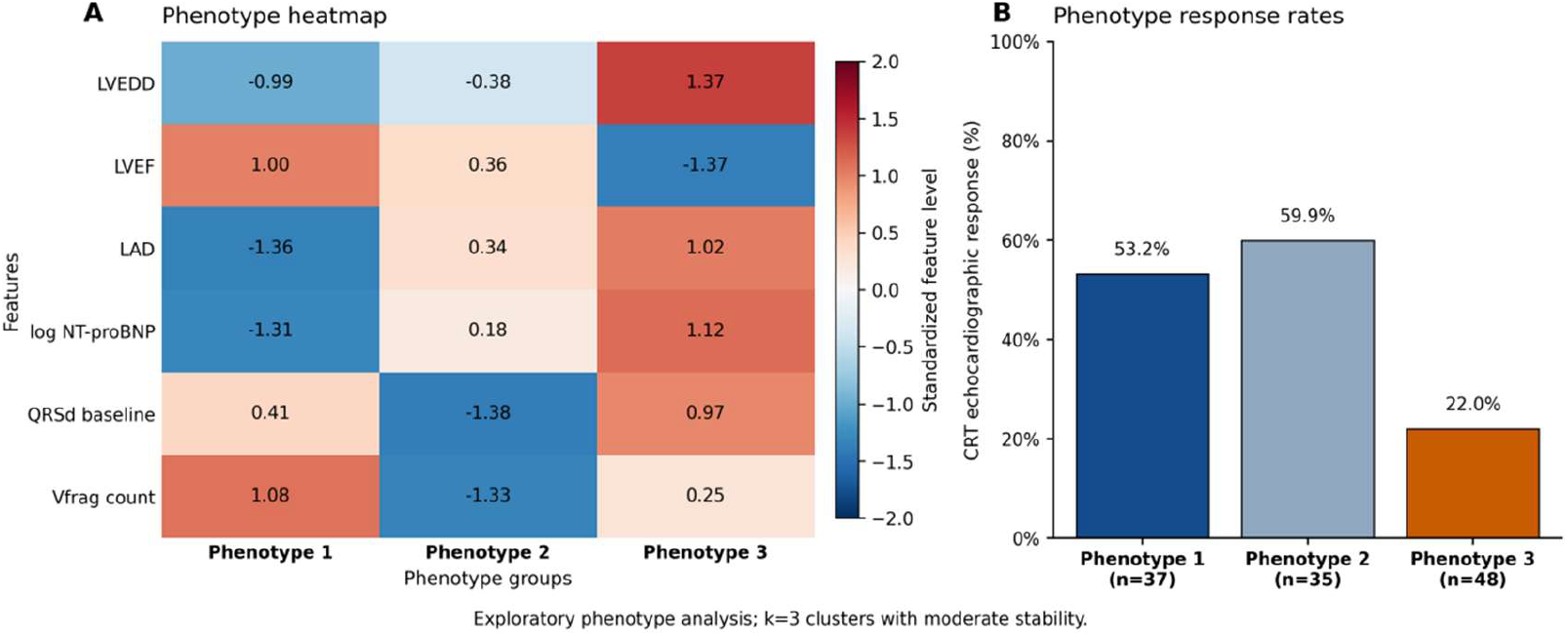
Exploratory phenotype profiles and echocardiographic response rates. (A) Heatmap showing standardized feature profiles of the three exploratory phenotype groups derived from pre-implant variables. (B) Mean 1-year echocardiographic CRT response rates across phenotype groups.

### Sensitivity analyses

Prespecified sensitivity analyses did not materially alter the main findings (Table 5). Complete-case analysis yielded directionally consistent performance compared with the multiply imputed analysis. Replacing Vfrag count with individual V1–V6 fragmentation indicators did not provide a material advantage over the more parsimonious summary measure. Removal of the posterolateral BiV LV lead-location indicator from the enhanced model did not improve model performance, supporting the limited retained value of procedural extension variables. Recalibration comparisons supported the intercept-recalibrated primary model as the preferred reporting version. An exploratory era-based analysis suggested a descriptive temporal gradient in response rates, but this did not alter model positioning.

**Table 5.** Summary of prespecified sensitivity analyses.

| Sensitivity analysis | Alternative specification | Key result | Interpretation | Main conclusion affected? |
| --- | --- | --- | --- | --- |
| Complete-case analysis | Fixed primary model | AUC 0.840; Brier 0.159; | Complete-case | No |
|  | refitted in complete-case cohort | calibration slope 1.000. MI apparent AUC 0.811; Brier 0.173. | findings were directionally consistent with the multiply imputed analysis. |  |
| Alternative fragmentation representation | Individual V1–V6 fragmentation indicators replaced Vfrag count | AUC 0.830; Brier 0.166; calibration slope 1.000. | Lead-specific indicators did not provide a material advantage over the more parsimonious Vfrag count. | No |
| Enhanced model without BiV lead-position indicator | Enhanced model refitted after removing the posterolateral BiV LV lead-location indicator | AUC 0.819; Brier 0.170; calibration slope 1.000. | Removing the position indicator did not meaningfully improve model performance; procedural extension remained limited. | No |
| Recalibration strategy comparison | Original, intercept-recalibrated, and shrunk primary model versions compared | Original/intercept-recalibrated AUC 0.820; Brier 0.169; slope 1.104. Shrunk AUC 0.819; Brier 0.172; slope 1.432. | Intercept-recalibrated primary model remained the preferred reporting version; shrinkage did not improve overall balance. | No |
| Era/temporal sensitivity | Early versus late implantation period by implantation order | Early era: n=60, response 48.3%; late era: n=60, response 36.7%. Mean LVEDD 57.3 vs 58.6 mm; mean log NT-proBNP 7.33 vs 7.25. | A temporal gradient was noted but was descriptive and insufficient to alter model positioning. | No; exploratory only |

Values are summarized from prespecified sensitivity analyses. AUC denotes area under the receiver operating characteristic curve. Brier score denotes overall prediction error. MI denotes multiple imputation. Vfrag count denotes the number of fragmented precordial leads among V1–V6. BiV denotes biventricular pacing; LV denotes left ventricular. The era/temporal analysis was exploratory and interpreted descriptively.

## Discussion

In this study of patients with non-LBBB heart failure undergoing CRT, baseline substrate provided a more stable framework for estimating 1-year echocardiographic response than procedural information alone. The value of this approach lies not in adding model complexity, but in integrating the structural, biomarker, and electrocardiographic domains in which CRT is delivered. Procedural variables remained clinically relevant, but their interpretation appeared dependent on the underlying substrate rather than on pacing strategy as an isolated label. This distinction is particularly important in non-LBBB patients, in whom conventional electrocardiographic eligibility criteria do not consistently identify a uniform resynchronization substrate^[7,8]^.

The clinical uncertainty addressed by this study stems from the fact that non-LBBB is an umbrella electrocardiographic category rather than a single disease mechanism. Contemporary evidence suggests that clinically relevant differences may be obscured when right bundle branch block, intraventricular conduction delay, and other non-LBBB morphologies are analyzed together. Patient-level evidence from randomized CRT trials has shown that CRT benefit differs across non-LBBB morphologies, with more favorable outcomes in intraventricular conduction delay than in right bundle branch block, challenging the practice of treating non-LBBB as a single category. A wide QRS complex outside typical LBBB does not necessarily indicate the same delayed left ventricular activation pattern or reversible mechanical substrate that underlies the most predictable response to conventional biventricular CRT^[8,20]^. The relatively low response rate in the present cohort is therefore clinically informative rather than unexpected; it is consistent with the uncertainty of CRT benefit in non-LBBB populations and reinforces the need for pre-implant substrate-based risk stratification rather than reliance on conduction morphology alone.

These findings also support a broader interpretation of CRT response in heart failure: electrical criteria are necessary but may not be sufficient to characterize response potential. Ventricular size, systolic function, atrial remodeling, biomarker burden, and pulmonary pressure each reflect different aspects of myocardial reserve, filling pressure, and chronic hemodynamic stress. Baseline QRS duration and fragmented QRS burden add a complementary electrical dimension, but neither alone captures whether the myocardium remains capable of reverse remodeling. Thus, the role of a substrate-based model is not to replace electrocardiographic selection, but to contextualize it within the structural and biological state of the patient. This framing is consistent with contemporary prediction and phenotyping work suggesting that CRT response is better understood as a multidimensional phenomenon than as a binary consequence of QRS morphology^[18-20]^.

The procedural implications should be interpreted within the rapidly evolving evidence comparing conduction system pacing with conventional biventricular CRT. Conventional BiV-CRT remains a foundational resynchronization strategy, whereas LBBAP and related conduction system pacing approaches have gained momentum because they may recruit the His–Purkinje system and achieve more physiologic ventricular activation. Recent randomized evidence has made this field more dynamic but not simpler^[9-14]^. The HeartSync-LBBP randomized trial suggested potential clinical advantages of left bundle branch pacing over biventricular pacing in selected patients with LBBB and severely reduced LVEF^[15]^. In contrast, the LEFT-BUNDLE-CRT trial did not establish LBBAP-CRT as noninferior to BiV-CRT in the intention-to-treat analysis of CRT-eligible patients with LBBB, although on-treatment analyses suggested more comparable efficacy^[16]^. These studies were not conducted in a non-LBBB population, but together they illustrate that the procedural question is unlikely to be resolved by a universal “LBBAP vs BiV” framing.

Against this background, the present study provides a complementary perspective focused on non-LBBB patients. It does not compare BiV pacing and LBBAP as causal treatment assignments, and it should not be interpreted as evidence that procedural technique is unimportant. Rather, it suggests that procedural labels alone may be insufficient to explain echocardiographic response once baseline substrate is considered. In clinical terms, this supports a substrate-first approach: before asking which pacing strategy is preferable, it may be necessary to ask whether the patient’s structural and electrical substrate is compatible with meaningful recovery. Future studies comparing BiV pacing, LBBAP, or hybrid resynchronization strategies may therefore be more informative if they stratify or enrich non-LBBB patients according to baseline substrate rather than treating conduction morphology alone as the primary selection framework.

The exploratory phenotype analysis further supports the view that non-LBBB CRT recipients are clinically heterogeneous. This analysis should be interpreted as heterogeneity mapping rather than as the discovery of fixed biological subtypes. Phenomapping approaches in heart failure and CRT can identify clinically interpretable patterns across multidimensional data, but the stability and transportability of such groups depend on the variables entered, cohort composition, and clustering strategy^[17,21,22]^. In the present study, the phenotype patterns separated patients along axes of structural remodeling, biomarker burden, atrial size, QRS duration, and QRS fragmentation. Given the moderate stability and single-center sample size, these groups should be considered hypothesis-generating. Their main value is to suggest that non-LBBB CRT response may be better studied within substrate-defined patterns than across the entire non-LBBB category.

Several limitations should be considered. This was a single-center retrospective study with a modest sample size, and the internally validated model requires external validation before clinical implementation. The primary endpoint was echocardiographic response rather than hard clinical outcomes such as heart failure hospitalization or mortality. LV end-systolic volume–based reverse remodeling was not available as the primary response definition. Procedural classification was based on the final successfully delivered pacing strategy documented at implantation. Although failed intended LBBAP without successful alternative CRT delivery was excluded, detailed electrophysiological metrics allowing uniform distinction between selective and nonselective left bundle branch capture were not available in all cases; therefore, LBBAP was analyzed as a clinical procedural strategy rather than as a granular capture subtype. The exploratory phenotype analysis was limited by sample size and moderate stability and should be viewed as hypothesis-generating^[26,27]^.

## Conclusion

In patients with non-LBBB heart failure undergoing CRT, baseline structural, biomarker, and electrocardiographic substrate provided the most stable framework for predicting 1-year echocardiographic response. Procedural variables, including pacing strategy and lead-position characteristics, added limited retained predictive value after internal validation, suggesting that implantation strategy should be interpreted alongside baseline substrate rather than as an isolated determinant of response. Exploratory phenotyping highlighted clinically relevant heterogeneity within the non-LBBB population and may inform future phenotype-guided studies of BiV pacing, LBBAP, and hybrid CRT strategies. External validation in larger multicenter cohorts with clinical outcome endpoints is needed before clinical implementation.

## Data Availability

The data that support the findings of this study are not publicly available because of institutional privacy and ethical restrictions. De-identified data may be made available from the corresponding author upon reasonable request and subject to approval by the relevant institutional authorities.

## Acknowledgments

The authors thank all members of the clinical electrophysiology, echocardiography, and device follow-up teams involved in the care and follow-up of the patients included in this study.

## Ethics Approval

This study was conducted in accordance with the Declaration of Helsinki and was approved by the Ethics Committee of Northern Jiangsu People’s Hospital, Yangzhou, China (approval number: 2021ky007). Given the retrospective design and use of de-identified clinical data, the requirement for written informed consent was waived by the ethics committee.

## Supplemental Material

Supplementary Methods

Tables S1–S6

Figures S1–S2

